# Benchmarking genome-wide association study causal gene prioritization for drug discovery

**DOI:** 10.1101/2025.09.23.25336370

**Authors:** Caleb Ji, Isabel Kerrebijn, Keon Arbabi, Marijn Schipper, Michael Wainberg

**Affiliations:** Lunenfeld-Tanenbaum Research Institute, Mount Sinai Hospital, Toronto, Canada; Department of Computer Science, University of Toronto, Toronto, Canada; Institute of Medical Science, University of Toronto, Toronto, Canada; Krembil Centre for Neuroinformatics, Centre for Addiction and Mental Health, Toronto, Canada; Department of Complex Trait Genetics, Center for Neurogenomics and Cognitive Research, Amsterdam Neuroscience, Vrije Universiteit Amsterdam, Amsterdam, The Netherlands; Department of Psychiatry, University of Toronto, Toronto, Canada; Division of Biostatistics, Dalla Lana School of Public Health, University of Toronto, Toronto, Canada

## Abstract

Drug discovery is costly, with billions spent on failed trials. Drugs with genetic support from genome-wide association studies (GWAS) have substantially greater odds of success, but how best to use GWAS to prioritize drug targets remains unclear. We evaluated the performance of GWAS causal gene prioritization methods from the Open Targets consortium by cross-referencing their predictions with drug trial outcomes for 445 diseases. We found that neither expression quantitative trait locus colocalization nor Open Targets’ locus-to-gene (L2G) score outperformed the simple nearest gene method at prioritizing which genes would become approved drug targets. Our findings inform the use of genetic evidence in drug discovery.

## Introduction

Drug development is an error-prone economic sinkhole, with up to 90% of investigational drugs failing in clinical trials^1,2^. In 2018, fewer than one new drug was approved per one billion USD spent, with failed candidates accounting for 60% of total development costs^3^. However, the cost efficiency of drug development may start to improve for the first time since 1950, partly due to advances in human genetics^3^. Recent analyses have demonstrated that drug development backed by human genetic evidence has a 2-3 fold higher success rate^4,5^. One major source of human genetic evidence is genome-wide association studies (GWAS), with over 7000 publications included in the GWAS Catalog as of 2025.^6^ The sheer scale of GWAS studies has provided hundreds of thousands of associations between complex traits and genetic variants, but around 90% of these variants map to non-coding regions, contributing to a lack of confidence in the causal genes underlying these associations.^7^ This uncertainty in causal gene identification may explain why GWAS shows lower predictive accuracy for drug target identification compared to Mendelian disease evidence, where the causal gene is unambiguously known^4,5^. While statistical approaches like colocalization with molecular quantitative trait loci (QTLs) and machine learning methods have emerged to address this challenge, their ability to identify therapeutically relevant targets remains largely unvalidated. Here, we address this gap by developing a benchmarking framework that uses drug clinical trial outcomes as a practical endpoint for assessing causal gene prioritization methods.

## Results

To benchmark causal gene prioritization methods against therapeutic outcomes, we integrated two complementary datasets. First, we sourced monotherapy clinical trial outcomes from Citeline Pharmaprojects, as curated by Minikel et al.^4^. After quality control, and the removal of active drug programmes (which count as neither successes nor failures, since they are still in progress), this dataset provided 14,958 target-indication pairs, each defined by a human gene target, a Medical Subject Headings (MeSH) indication, and the maximum clinical trial phase achieved (Phase 1, Phase 2, Phase 3, or preclinical). Second, we leveraged the Open Targets Platform^8^, filtering the >100,000 GWAS with available summary statistics to a final set of 445 GWAS, each of which was the best-powered GWAS available for a particular disease with a MeSH ID in Pharmaprojects (**Figure 1**, Methods). Integrating the clinical and genetic data required harmonizing their disparate disease ontologies: MeSH and Experimental Factor Ontology (EFO), respectively. We used an ensemble mapping approach that ensured precision while maintaining broad coverage (Methods).

**Fig. 1:**
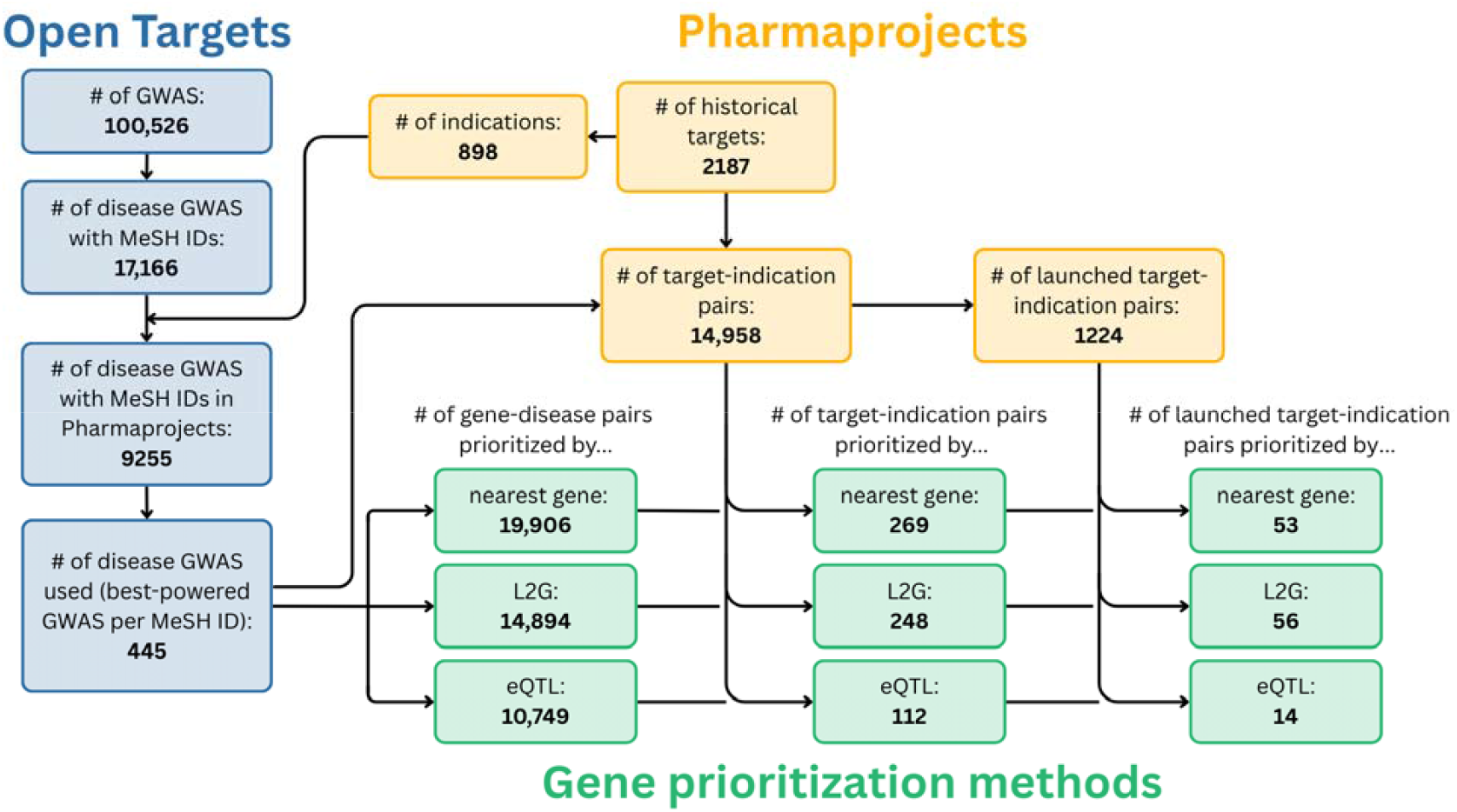
Summary of data processing. Candidate causal genes prioritized from disease GWAS by the Open Targets consortium were cross-referenced against the approval status of drugs targeting the same genes and disease indications in the Pharmaprojects database. Diseases were defined according to Medical Subject Headings (MeSH) IDs. Three causal gene prioritization strategies were compared: nearest gene, Open Targets’ machine learning-based locus-to-gene (L2G) score, and eQTL colocalization.

We evaluated three causal gene prioritization methods readily available from Open Targets: expression quantitative trait locus (eQTL) colocalization, the machine learning-based locus-to-gene (L2G) score developed by Open Targets^9^, and the nearest gene method (Methods). We evaluated each method’s ability to predict clinical success by comparing the targets of launched drugs to those that failed during development, i.e. targets with a historical but no active clinical trial phase. For each method, we quantified success rates using odds ratios (OR) with 95% confidence intervals (CI), relative to a baseline of drug targets lacking that method’s genetic evidence.

We found that the L2G score (OR = 3.14, 95% CI of 2.31 to 4.28; **Figure 2a**) and the nearest gene method (OR = 3.08, 95% CI of 2.25 to 4.11) were similarly predictive of drug success, while eQTL colocalization did not significantly predict drug approval (OR = 1.61, 95% CI of 0.92 to 2.83). Since over two-thirds of genes prioritized by eQTL colocalization were nearest genes (77 of 112, 68.75%), we next tested whether eQTLs provide value when they disagree with the nearest gene method. When removing nearest gene predictions, eQTL colocalization was associated with an even lower likelihood of approval (OR = 0.33, 95% CI of 0.05 to 2.41), identifying only one launched drug target out of thirty-five prioritized targets.

**Fig. 2:**
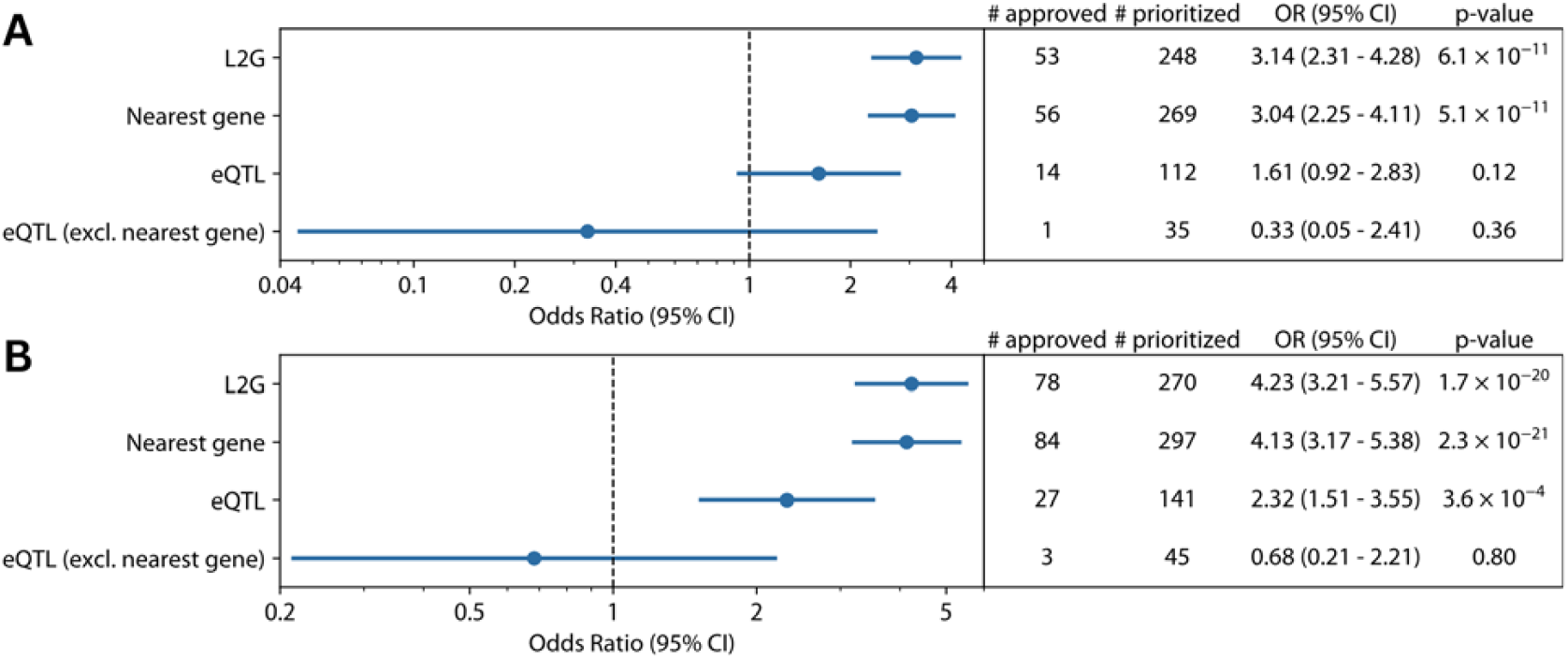
Enrichment of causal gene prioritization strategies for successful drug targets. Enrichment of genes prioritized by various strategies among **A)** launched target-indication pairs and **B)** launched target-specialty pairs, relative to those without genetic evidence. “Nearest gene” refers to the gen closest to the lead variant. “L2G” denotes the gene at the locus with a locus-to-gene (L2G) score ≥□75% of the sum of the L2G scores across all genes at the locus, if any. “eQTL” refers to genes with eQTL colocalizations according to coloc (PPH4 ≥□0.8) or eCAVIAR (CLPP ≥□0.1; Methods). Odds ratios and 95% confidence intervals (CI) are given for each genetic evidence source, with statistical significance assessed using Fisher’s exact test. “# prioritized” denotes the number of target-indication pairs prioritized by each method, and “# approved” denotes the number of these that resulted in launched drugs.

One potential limitation of this analysis is the recurrence of drug targets across related indications. For example, drugs targeting DRD2 are launched for many psychiatric indications, such as anxiety disorders, depression, bipolar disorder and schizophrenia. Getting a drug approved for one indication makes it easier to get approved for other indications, because clinical safety has already been shown and information on the drug’s minimal anticipated biological effect level (MABEL) and no-observed-adverse-effect level (NOAEL) is available. To address this, we performed a secondary analysis that grouped indications by clinical specialty into target-specialty pairs (Methods), based on the premise that most such repurposings will be within the same specialty, while cross-specialty repurposings are far less trivial and still deserve to be treated separately. This specialty-level analysis increased all methods’ odds ratios, since it counts cases where a target fails for one indication, but is later launched for a different indication, as successes rather than failures. However, this analysis maintained the relative performance of the L2G score (OR = 4.23, 95% CI of 3.21 to 5.57; **Figure 2b**), the nearest gene method (OR = 4.13, 95% CI of 3.17 to 5.38), and eQTL colocalization (OR = 2.32, 95% CI of 1.51 to 3.55). eQTL colocalization was once again associated with a lower likelihood of approval after removing nearest gene predictions (OR = 0.68, 95% CI of 0.21 to 2.21), confirming its limited independent predictive value.

Finally, we tested whether eQTL colocalization or L2G provide additional value for nearest genes, which as noted above are associated with greater odds of drug approval (OR = 3.04, 95% CI of 2.25 to 4.11; **Figure S1**). L2G performed similarly to the nearest-gene heuristic alone (OR = 3.27, 95% CI 2.39 to 4.49). In contrast, eQTL colocalization was associated with a lower odds ratio (OR = 2.29, 95% CI 1.26 to 4.17). This suggests that within the set of nearest genes, those with additional eQTL support are less likely to be approved than those without, mirroring the negative trend observed when taking out nearest genes.

## Discussion

Our analysis provides a systematic evaluation of causal gene prioritization methods against pharmaceutical clinical trials, a practical and comprehensive benchmark for target validation. While individual drugs may fail for reasons unrelated to target selection, systematic patterns of success and failure across many programs should tend to reflect underlying target validity. This benchmark is also practically relevant, as it directly measures the success of translating genetic insights into approved therapeutics. Our approach aims to be robust to the fact that pharmaceutical companies are biased in which targets they pursue, based on prior approvals, genetic evidence, and other factors. Our core result is that neither expression quantitative trait locus colocalization nor the machine learning-based locus-to-gene (L2G) score improved upon the performance of the much simpler nearest gene method at prioritizing approved drug targets.

Several factors may explain the inability of eQTL colocalization to outperform the nearest-gene baseline. GWAS and eQTL studies are systematically biased towards discovering different types of genes. GWAS signals are enriched near functionally complex and evolutionarily constrained genes, often in distal enhancer regions, whereas detectable eQTLs are depleted from these genes and cluster strongly near the transcription start sites of simpler, less constrained genes. This is likely a consequence of natural selection, which purges large-effect eQTLs at functionally critical genes, rendering them less visible to eQTL studies, whereas GWAS more readily detects signals at these genes^10^. An analysis of genes in which coding variants have phenotypic effects, identified via familial studies of Mendelian disorders or exome-wide burden tests, showed that only about 8% also showed evidence of eQTL colocalization with the corresponding GWAS trait^11^, suggesting a “missing regulation” problem in which many non-coding associations act through regulatory mechanisms that are context-specific or dynamic, and therefore not detectable via homeostatic eQTLs. Similarly, an analysis of metabolomics GWAS, where causal genes can often be inferred from metabolic pathway knowledge, demonstrated a ∼75% false-positive rate for cis-eQTL evidence, whereas the nearest gene to the sentinel variant provided a much stronger enrichment for true causal genes^12^. While Open Targets’ broad, pan-tissue approach to eQTL colocalization may have contributed to higher false positive rate, this prior work suggests that tissue-specific analyses would not fully address the underperformance of eQTL colocalization.

Our focus on established causal gene prioritization methods from Open Targets is a key feature of this study. This approach is distinct from, and complementary to, recent benchmarks of network-based methods, which prioritize targets based on their interaction with known disease genes rather than by resolving specific GWAS signals^13^. A limitation of this focus is that it excludes emerging state-of-the-art methods. For instance, while L2G achieved the highest enrichment of the three methods tested, its negligible improvement over the simpler nearest gene method might be surpassed by more recent machine learning methods, such as FLAMES^14^ and methods based on large language models^15^. Future work should determine whether these approaches’ reported gains in predictive accuracy translate to better prediction of successful drug targets.

These findings provide insight into widely used approaches to GWAS causal gene prioritization and highlight opportunities for improvement. Continued advances in methods for identifying causal genes from GWAS are essential because human genetics offers enormous unrealized opportunities for drug development. Active drug development programmes are only slightly more enriched for genetic evidence than historical programmes,^4^ and analysis of publicly available clinical trial data suggests that almost 60% of disease areas with established GWAS associations lack corresponding drug development activity^16^. Enhancing causal gene prioritization methods promises to improve the accuracy of interpreting GWAS findings and reduce the cost of drug discovery and development.

## Methods

### Drug development outcome data

Curated drug development data were obtained from Minikel et al. (2024)^17^, originally sourced from Citeline’s Pharmaprojects^18^ (API access date: Dec 22, 2022). From this dataset, we extracted target-indication pairs and the maximum clinical trial phase achieved for each pair. We filtered out duplicate entries and those with missing indications. A target was defined as the gene encoding the protein modified by the drug, and an indication was defined by its Medical Subject Headings (MeSH)^19^ unique identifier. For all analyses, a “successful” outcome was defined as a target-indication or target-specialty pair corresponding to a drug that has been launched, while a “failure” was defined as a target-indication or target-specialty pair corresponding to a historical drug program that did not result in a launched drug. We removed any ‘active’ (drug development ongoing) target-indication pairs from the analysis, since they do not conclusively represent either a success or a failure.

### Genetic evidence from the Open Targets Platform

Genetic evidence data were sourced from the Open Targets Platform (version 25.06)^8^. We downloaded the GWAS Study, Credible Set, Locus-to-gene (L2G) prediction, and colocalization (COLOC and eCaviar) datasets. Both COLOC and eCaviar analyses utilize Open Targets’ curated expression Quantitative Trait Locus (eQTL) datasets, which solely contain cis-eQTLs (local expression effects within the same chromosomal region as the variant).

### Data processing and integration

Open Targets GWAS studies with multiple Experimental Factor Ontology (EFO)^20^ identifiers were filtered out, as they usually represented conditional, combined, or complex phenotypes. GWAS for traits with names that included the words “MTAG”, “multivariate analysis”, or “medication” were also removed as they are imperfect proxies for the corresponding disease.

To align indications between the drug development and genetic datasets, we mapped the EFO identifiers from GWAS studies to MeSH identifiers. We harmonized terms using a union of four approaches: (1) direct MeSH cross-references from the EFO OWL database; (2) Unified Medical Language System (UMLS)^21^ cross-references from EFO, subsequently mapped to MeSH using the UMLS database; (3) exact (case-insensitive) string matching of the GWAS trait name to MeSH terms and synonyms (“entry terms”); and (4) exact (case-insensitive) string matching of the GWAS trait name to UMLS terms, which were then mapped to MeSH.

Following the mapping, multiple GWAS studies often corresponded to a single MeSH indication. To reduce redundancy and select a high-quality representative study for each indication, we selected the single GWAS with the highest number of credible sets as a measure of statistical power, breaking ties on the number of samples. Further ties were broken through manually choosing the most relevant GWAS based on the study. All subsequent analyses were subset to these selected GWAS. Indications that did not have a GWAS were not included in the analysis.

### Causal gene prioritization methods

Locus-to-gene (L2G)^9^ is a method developed by Open Targets to nominate candidate causal genes at GWAS risk loci. It scores genes based on variant-to-gene evidence along three main categories: distance, QTL colocalization, and Variant Effect Predictor (VEP) annotations^22^. The model weights were calculated through training on manually curated ‘gold-standard’ causal genes from pharmacological and genetic evidence. L2G predictions were filtered to genes scoring ≥ 75% of the sum of L2G scores for all genes in the locus, a threshold with highest enrichment in previous studies^4^. Odds ratios were nearly identical when using a threshold of 50% instead of 75%, suggesting the precise threshold does not affect performance much. COLOC^23^ is a Bayesian colocalization method chosen by Open Targets that uses variant-level Bayes factors from statistical fine-mapping to derive the posterior probability of four hypotheses, where hypothesis 4 (H4) measures the probability that GWAS and eQTL signals share the same causal variant. COLOC prediction was thresholded on a posterior probability of H4 ≥ 0.8.

eCaviar^24^ is a heuristic colocalization method used by Open Targets when variant-level Bayes factors were unavailable. It calculates a locus’s colocalization posterior probability (CLPP) by multiplying each variant’s posterior probability from fine-mapping the GWAS with its posterior probability from fine-mapping the eQTL study, and then summing these products across the locus. eCaviar prediction was thresholded on a CLPP score ≥ 0.1. Since Open Targets used eCaviar and COLOC at mutually exclusive loci based on the availability of variant-level Bayes factors, we combined their predictions into a single eQTL colocalization predictor.

Nearest gene selection prioritized gene bodies closest to the GWAS variant with the lowest p-value (lead variant). The gene body was defined to start at the transcription start site and end at the transcription end site using gencode version 46. If two genes had the same distance to the lead variant, both were included as nearest genes.

### Specialty analysis

MeSH terms were categorized into 18 therapeutic specialties following the categorization scheme of Minikel et al.^4^. When indications mapped to multiple specialties, all specialties were included except for two circumstances: cancer indications were assigned exclusively to the cancer specialty, and “other” was used only when no specific specialty was applied. For each target-specialty combination, we defined the maximum clinical phase as the highest phase reached by any constituent target-indication pair. Similarly, a causal gene prioritization method was considered to support a target-specialty pair if it provided genetic evidence for any target-indication pair within that group.

### Statistical analysis

For each prioritization method, we constructed a 2 × 2 contingency table based on whether a target-indication pair was prioritized by the method and whether it resulted in a launched drug. True Positives (denoted “# approved” in the figures) were target-indication pairs prioritized by the genetic evidence method that resulted in a launched drug. Predicted Positives (“# prioritized”) were all target-indication pairs prioritized by the genetic evidence method. From this table, we calculated the odds ratio (OR) and a Fisher exact p-value to quantify the enrichment of successfully launched drugs among the methods for genetic support.

## Data Availability

All data produced in the present study are available upon reasonable request to the authors

---

**Supplementary Fig. 1:**
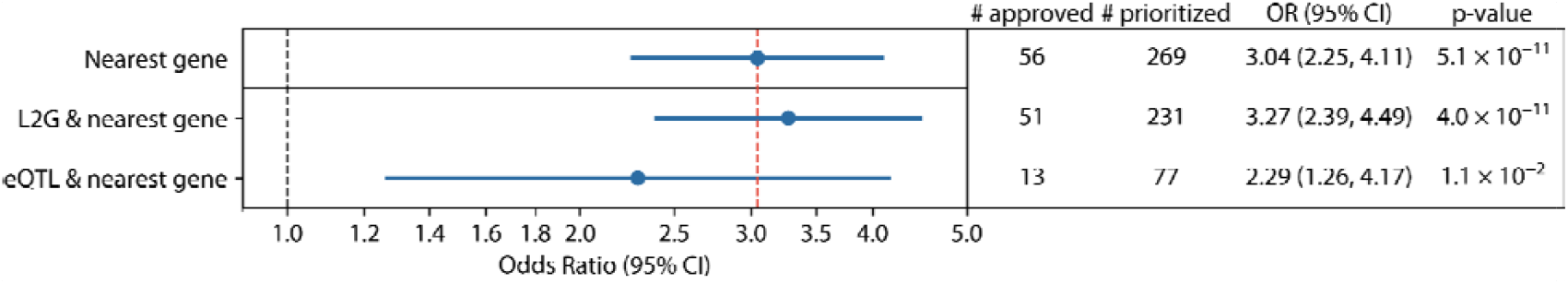
Enrichment of gene prioritization strategies for successful drug targets among nearest genes. Enrichment of genes prioritized by L2G and eQTL, restricted to those also identified by the nearest-gene method, among launched target-indication pairs. See Fig. 2 caption for definitions and statistical details.

## Supplementary Data

**Supplementary Data 1: GWAS selection details**

*The 471 GWAS retained after filtering for the best-powered disease GWAS with Medical Subject Headings (MeSH) IDs from Pharmaprojects. Credible sets and sample size are shown as indicators of study power, which were used for ranking and filtering. The source GWAS name and corresponding MeSH ID are included to document the disease-mapping scheme*.

**Supplementary Data 2: Target indication summary**

*The 14,958 target-indication pairs and their maximum clinical phase attained. MeSH term mappings to specialties included*.

## Notes

### Competing Interest Statement

The authors have declared no competing interest.

### Funding Statement

This study was funded by the Canadian Institutes of Health Research (CIHR MHP 192163; CIHR FBD 199459) and by the Natural Sciences and Engineering Research Council of Canada (NSERC) Undergraduate Student Research Award (URSA).

